# Genomic epidemiology of a densely sampled COVID-19 outbreak in China

**DOI:** 10.1101/2020.03.09.20033365

**Authors:** Lily Geidelberg, Olivia Boyd, David Jorgensen, Igor Siveroni, Fabricia F. Nascimento, Robert Johnson, Manon Ragonnet-Cronin, Han Fu, Haowei Wang, Xiaoyue Xi, Wei Chen, Dehui Liu, Yingying Chen, Mengmeng Tian, Wei Tan, Junjie Zai, Wanying Sun, Jiandong Li, Junhua Li, Erik M Volz, Xingguang Li, Qing Nie

## Abstract

Analysis of genetic sequence data from the SARS-CoV-2 pandemic can provide insights into epidemic origins, worldwide dispersal, and epidemiological history. With few exceptions, genomic epidemiological analysis has focused on geographically distributed data sets with few isolates in any given location. Here we report an analysis of 20 whole SARS-CoV 2 genomes from a single relatively small and geographically constrained outbreak in Weifang, People’s Republic of China. Using Bayesian model-based phylodynamic methods, we estimate a mean basic reproduction number (***R***_0_) of 3.47 (95% highest posterior density interval: 1.78-5.47) in Weifang, and a mean effective reproduction number (***R****_t_*) that falls below 1 on February 2nd. We further estimate the number of infections through time and compare these estimates to confirmed diagnoses by the Weifang Centers for Disease Control. We find that these estimates are consistent with reported cases and there is unlikely to be a large undiagnosed burden of infection over the period we studied.

## Introduction

We report a genomic epidemiological analysis of one of the first geographically concentrated community transmission samples of SARS-CoV 2 genetic sequences collected outside of the initial outbreak in Wuhan, China. These data comprise 20 whole genome sequences from confirmed COVID-19 cases in Weifang, Shandong Province, People’s Republic of China. The data were collected over the course of several weeks up to February 10, 2020 and overlap with a period of intensifying public health and social distancing measures. These interventions included public health messaging, establishing phone hotlines, encouraging home isolation for recent visitors from Wuhan (January 23-26), optimising triage of suspected cases in hospitals (January 24), travel restrictions (January 26), extending school closures, and establishing ‘fever clinics’ for consultation and diagnosis (January 27) (***Mao, 2020***). Phylodynamic analysis allows us to evaluate epidemiological trends after seeding events which took place in mid to late January, 2020.

The objective of our analysis is to evaluate epidemiological trends based on national surveillance and response efforts by Weifang Centers for Disease Control (CDC). This analysis provides an estimate of the initial rate of spread and reproduction number in Weifang City. In contrast to the early spread of COVID-19 in Hubei Province of China, most community transmissions within Weifang took place after public health interventions and social distancing measures were put in place. We therefore hypothesise that genetic data should reflect a lower growth rate and reproduction number than was observed in Wuhan, which decreases over time. A secondary aim is to estimate the total numbers infected and to evaluate the possibility that there is a large unmeasured burden of infection due to imperfect case ascertainment and a large proportion of infections with mild or asymptomatic illness.

## Methods and Materials

### Epidemiological investigation, sampling, and genetic sequencing

As of 10 February 2020, 136 suspected cases, and 214 close contacts were diagnosed by Weifang Center for Disease Control and Prevention. 28 cases were detected positive with SARS-CoV-2. Viral RNA was extracted using Maxwell 16 Viral Total Nucleic Acid Purification Kit (Promega AS1150) by magnetic bead method and RNeasy Mini Kit (QIAGEN 74104) by column method. Quantitative reverse transcription polymerase chain reaction (RT-qPCR) was carried out using 2019 novel coronavirus nucleic acid detection kit (BioGerm, Shanghai, China) to confirm the presence of SARS-CoV-2 viral RNA with cycle threshold (Ct) values range from 17 to 37, targeting the high conservative region (ORF1ab/N gene) in SARS-CoV-2 genome.

Metagenomic sequencing: The concentration of RNA samples was measurement by Qubit RNA HS Assay Kit (Thermo Fisher Scientific, Waltham, MA, USA). The enzyme DNase was used to remove host DNA. The remaining RNA was used to construct the single-stranded circular DNA library with MGIEasy RNA Library preparation reagent set (MGI, Shenzhen, China). Purified RNA was then fragmented. Using these short fragments as templates, random hexamers were used to synthesize the first-strand cDNA, followed by the second strand synthesis. Using the short double-strand DNA, a DNA library was constructed through end repair, adaptor ligation, and PCR amplification. PCR products were transformed into a single strand circular DNA library through DNA-denaturation and circularization. DNA nanoballs (DNBs) were generated with the single-stranded circular DNA library by rolling circle replication (RCR). The DNBs were loaded into the flow cell and pair-end 100bp sequencing on the DNBSEQ-T7 platform 8 (MGI, Shenzhen, China). 20 genomes were assembled with length from 26,840 to 29,882 nucleotides. The median age of patients was 36 (range:6-75). Two of twenty patients suffered severe or critical illness.

The Weifang sequences are deposited in GISAID (gisaid.org).

To analyse these sequences, we have adapted model-based phylodynamic methods which were previously used to estimate growth rates and reproduction numbers using sequence data from Wuhan and exported international cases (***Volz et al., 2020***).

### Mathematical model

The phylodynamic model is designed to account for non-linear epidemic dynamics in Weifang with a realistic course of infection (incubation and infectious periods), variance in transmission rates which can influence epidemic size estimates, and migration of lineages in and out of Weifang.

#### Nonlinear epidemiological dynamics in Weifang

The maximum number of daily confirmed COVID-19 cases occurred on February 5, but it is unknown when the maximum prevalence of infection occurred. To capture a nonlinear decrease in cases following epidemic peak, and to account for a realistic distribution of generation times, we use an extension of the susceptible-exposed-infectious-recovered (SEIR) model (***Keeling and Rohani, 2011***) for epidemic dynamics in Weifang, shown in Equations 1-5.

#### Variance in transmission rates

To estimate total numbers infected, the phylodynamic model must account for epidemiological variables which are known to significantly influence genetic diversity (***Lloyd-Smith et al., 2005***). Foremost among these is the variance in offspring distribution (number of transmissions per primary case). We draw on previous evidence based on the previous SARS epidemic which indicates that the offspring distribution is highly over-dispersed. High variance of transmission rates will reduce genetic diversity of a sample and failure to account for this factor will lead to highly biased estimates of epidemic size (***i et al., 2017***). Recent analyses of sequence data drawn primarily from Wuhan has found that high over-dispersion was required for estimated cases to be consistent with the epidemiological record (***Volz et al., 2020***). Models assuming low variance in transmission rates between people would generate estimates of cases that are lower than the known number of confirmed cases. Separately, Endo et al. (***Endo et al., 2020***) found that high over-dispersion is required to reconcile estimated reproduction numbers with the observed frequency of international outbreaks. We therefore elaborate the SEIR model to with an additional compartment ***J*** which has a higher transmission rate (*τ*-fold higher) than the ***I*** compartment.

The variance of the implied offspring distribution is calibrated to give similar overdispersion from the SARS epidemic.

Upon leaving the incubation period individuals progress to the ***J*** compartment with probability *p_h_*, or otherwise to ***I***. The model is implemented as a system of ordinary differential equations:

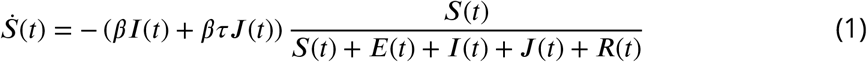

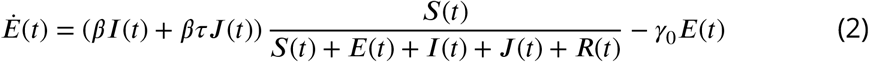

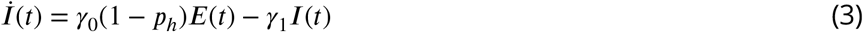

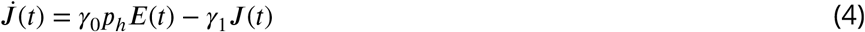

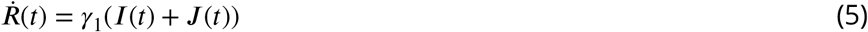

#### Importation of lineages from Wuhan

The outbreak in Weifang was seeded by multiple lineages imported at various times from the rest of China. We therefore account for location of sampling in our model. Migration is modelled as a bi-directional process with rates proportional to epidemic size in Weifang. The larger international reservoir of COVID-19 cases ***Y***(*t*) serves as a source of new infections and is assumed to be growing exponentially over this period of time.

The equation governing this population is

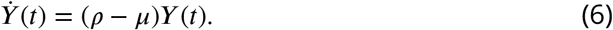

Migration only depends on the size of variables in the Weifang compartment and thus does not influence epidemic dynamics;it will only influence the inferred probability that a lineage resides within Weifang. Fora compartment *X* (E,I, or J), *η* is the per lineage rate of migration out of Weifang and the total rate of migration in and out of Weifang is *ηX*.

#### Model fitting

During phylodynamic model fitting *β* and *ρ* are estimated. Additionally, we estimate initial sizes of ***Y***, ***E***, and ***S***. Other parameters are fixed based on prior information. We fix 1/*γ*_0_ = 4.1 days and 1/*γ*_1_ = 3.8 days. We set *p_h_* = 0.20 and *τ* = 74 which yields a dispersion of the reproduction number that matches a negative binomial distribution with *k =* 0.22 if *R*_0_ = 2, similar to values estimated for the 2003 SARS epidemic (***Lloyd-Smith et al., 2005***).

### Phylogenetic analysis

We aligned the 20 Weifang sequences using MAFFT (***Katoh and Standley, 2013***) with a previous alignment of 50 non-identical SARS-CoV 2 sequences from outside of Weifang (***Volz et al., 2020***), provided by GISAID (***Elbe and Buckland-Merrett, 2017***).

Maximum likelihood analysis was carried using IQTree (***Minh et al., 2019***) with a HKY+G4 substitution model and a time-scaled tree was estimated using tree-dater 0.5.0 (***Volz and Frost, 2017***). Two outliers according to the molecular clock model were identified and removed using treedater which was also used to compute the root to tip regression.

Bayesian phylogenetic analysis was carried out using BEAST 2.6.1 (***Bouckaert et al., 2019***) using a HKY+G4 substitution model and a strict molecular clock. The phylodynamic model was implemented using the PhyDyn package v1.3.7 (***Volz and Siveroni, 2018***) using the QL likelihood approximation and the RKODE solver. The model was fitted by running 8 MCMC chains of 30 million steps in parallel, and combining chains after removing 50% burn-in. In order to demonstrate the added utility of the sequence data, the analysis was repeated assuming a constant likelihood, i.e. sampling only from the prior probability distributions.

The *ggtree* package was used for all phylogeny visualizations (***Yu et al., 2017***).

Code to replicate this analysis and BEAST XML files can be found at https://github.com/emvolz/weifang-sarscov2.

## Results

Despite an initial rapid increase in confirmed cases in Weifang in late January and early February, the number of confirmed cases by Weifang CDC show that the outbreak peaked quite early and maximum number of cases occurred on February 5. Phylodynamic analysis supports the interpretation that control efforts reduced epidemic growth rates and contributed to eventual control. ***Figure 1***A shows the estimated time scaled phylogeny (maximum clade credibility) including 20 lineages sampled from distinct patients in Weifang and 50 genomes sampled from Wuhan and internationally. ***Figure 1***B illustrates the phylodynamic model which was co-estimated with the phylogeny which provides estimates of epidemiological parameters summarised in ***Table 1***.

**Figure 1.**
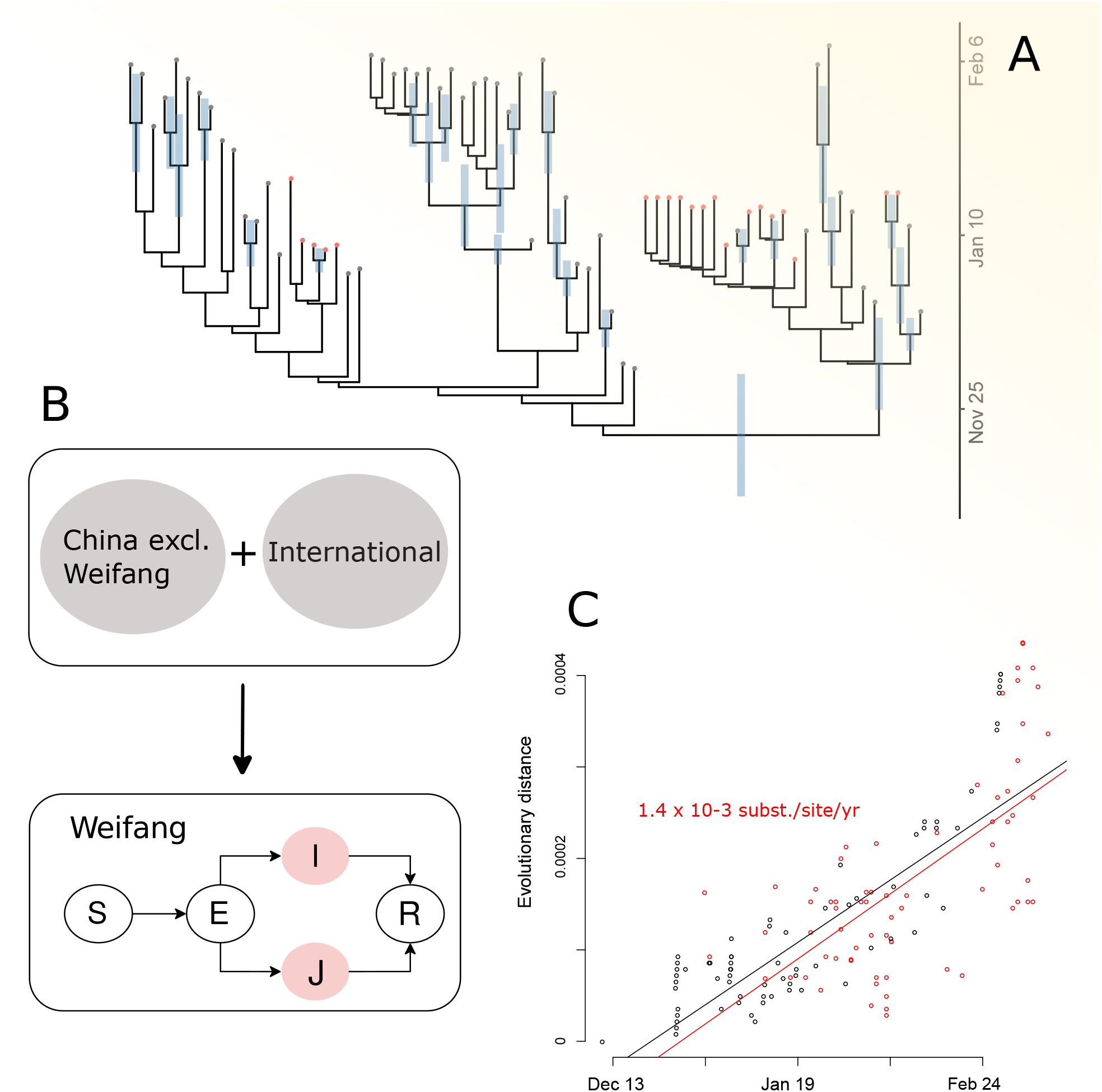
Phylodynamic estimates and epidemiological model. A. A time scaled phylogeny co-estimated with epidemiological parameters. Red and grey tips correspond to samples from inside and outside Weifang, China respectively. The credible interval of time to most recent common ancestor (TMRCA) is shown as a blue bar for all nodes with more than 50% posterior probability support. B. A diagram representing the structure of the epidemiological SEIR model which was fitted in tandem with the time scaled phylogeny. Colours correspond to the state of individuals sampled and represented in the tree (A). Note that infected and infectious individuals may occupy a low transmission state (I) or a high transmission rate state (J) to account for high dispersion of the reproduction number. C. A root to tip regression (red and black points indicate sample and internal nodes respectively) showing approximately linear increase in diversity with time of sampling. **Figure 1-Figure supplement 1**. Maximum likelihood time tree. **Figure 1-Figure supplement 2**. Tree posterior density plot. **Figure 1-Figure supplement 3**. Estimated posterior TMRCA for Weifang lineages.

The estimated cumulative and daily number of infections are shown in Figure ***Figure 2A*** and ***Figure 2B*** respectively. We estimate the peak of daily infections in late January, preceding the time series of confirmed cases by about a week;this is expected due to delays from infection to appearance of symptoms and delays from symptoms to diagnosis. The genetic data are strongly informative about timing and size of the epidemic peak: Trajectories sampled from the Bayesian prior distribution have a smaller and later epidemic peak (c.f. ***Figure 2)*** with much less precision. We also estimate that a maximum of 20% of infections were diagnosed; an unknown proportion of infections will be missed by the surveillance system due to very mild, sub-clinical, or asymptomatic infection. Our central estimate for the cumulative number infected on 10 February is 209 (HPD: 50-770), compared to 44 cumulative confirmed cases at the end of February. This supports the hypothesis that there was a modest (but not large) burden of infection in Weifang over the period that the sequence data were sampled.

**Figure 2.**
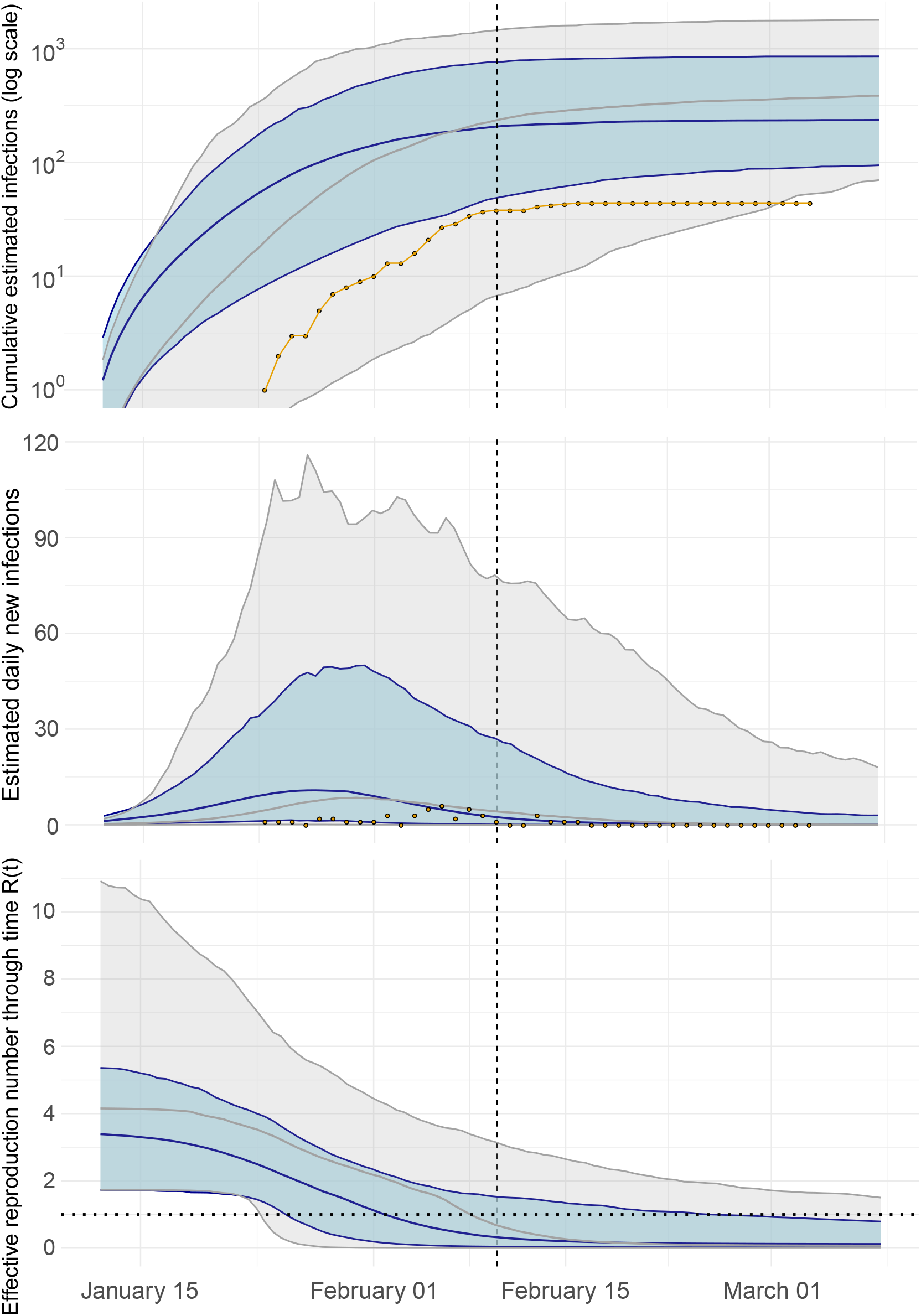
Epidemiological trajectory of the Weifang SARS-CoV-2 epidemic when fitting the SEIR model to genetic data (blue) and sampling only from prior (grey). Solid lines and shaded area reflect posterior median and 95% HPD. The vertical dashed line represents the date of the last sequence sampled in Weifang. A. Cumulative estimated infections through time compared to cumulative cases (yellow points) reported by Weifang CDC. B. Daily estimated infections through time compared to daily reported cases (yellow points). C. Effective reproduction number through time R(t). The horizontal dotted line indicates R(t) = 1. **Figure 2-Figure supplement 1**. Sample distribution through time. **Figure 2-Figure supplement 2**. Estimate frequency reported through time.

**Table 1.** Summary of primary epidemiological and evolutionary parameters, including Bayesian prior distributions and estimated posteriors. Posterior uncertainty is summarised using a 95% highest posterior density (HPD) interval.

| Parameter | Prior | Posterior mean | 95% HPD |
| --- | --- | --- | --- |
| Initial infected | Exponential(mean=1) | 4.4 | 0.73-9.9 |
| Initial susceptible | Exponential(mean=500) | 322 | 106-930 |
| Migration rate<sup>1</sup> | Exponential(mean=10) | 1.53 | 0.71-1.98 |
| Reproduction number | Log-normal(mean log=1.03, sd log=0.5) | 3.47 | 1.78-5.47 |
| Molecular clock rate<sup>2</sup> | Uniform(0.0007,0.003) | 0.0012 | 0.00086-0.0016 |
| Transition/transversion | Log-normal(mean log=1, sd log=1.25) | 7.2 | 4.4-11.4 |
| Gamma shape | Exponential(mean=1) | 0.32 | 0.0077-1.7 |

<sup>1</sup> Units: Migrations per lineage per year.
<sup>2</sup> Units: Substitutions per site per year.

Effective reproduction number over time is shown in Figure 2C. We estimate ***R***_0_ = 3.47 (95% HPD:1.78-5.47) and the initial growth rate in cases was approximately 22% per day, consistent with those estimated in other settings and during the early epidemic in Wuhan (***Alimohamadi et al., 2020***). Sampling from the prior yields a much higher estimate for ***R***_0_ with an unrealistic HPD upper bound over 10. We detect a significant decrease in effective reproduction number as the epidemic progressed, during a period (late January) when Weifang was implementing a variety of public health interventions and contact tracing to limit epidemic spread. Our central estimate of R(t) drops below 1 on the 2nd of February.

As well as providing novel epidemiological estimates, our results point to the significance of realistic modelling for fidelity of phylogenetic inference. The use of a model-based structured coalescent prior had large influence over estimated molecular clock rates and inferred time to most recent common ancestors (TM-201 RCAs).***Figure Supplement 1*** shows that maximum likelihood inference of time-scaled phylogenies produces a distribution of TMRCAs which are substantially different to the Bayesian model-based analysis. Choice of population genetic prior will have a large influence on phylogenetic inference based on sparse or poorly informative genetic sequence data. Among the 20 Weifang sequences included in this analysis, there is mean pairwise difference of only three single nucleotide polymorphisms and less than twice as much diversity observed among the remainder of the sequences we studied. There is correspondingly low confidence in tree topology (Figure Supplement 2), and only three monophyletic Weifang clades had greater than 50% posterior probability, none of which larger than three samples.

The earliest Weifang sequence was sampled on January25 from a patient who showed first symptoms on January 16. These dates cover a similar range as the posterior TMRCA of all Weifang sequences (Figure Supplement 3).

## Discussion

Our analysis of 20 SARS-CoV-2 genomes from Weifang, China has confirmed independent observations regarding the rate of spread and burden of infection in the city. Surveillance of COVID-19 is rendered difficult by high proportions of illness with mild severity and an unknown proportion of asymptomatic infection (***Guan et al., 2020***). The extent of under-reporting and case ascertainment rates has been widely debated. Analysis of genetic sequence data provides an alternative source of information about epidemic size. We do not find evidence for a large hidden burden of infection within Weifang, with an estimated total number of cases around 209 by the end of the outbreak.

Our decreasing central estimate of ***R****_t_* over time, falling below 1 on February 2nd, suggests a slower rate of spread outside of Wuhan and effective control strategies implemented in late January. It is consistent with a previous modelling study of Shandong province (Zhang et al., 2020), which showed that ***R****_t_* fell below 1 on January 29th. Our posterior molecular clock rate shown in ***Table 1*** is consistent with previous estimates of SARS-CoV-2 phylogenetic analyses (***Nie et al., 2020***).

While the value of pathogen genomic analysis is widely recognised for estimating dates of emergence (***Gire et al., 2014***) and identifying animal reservoirs (***Zhou et al., 2020; Dudas et al., 2018***), analysis of pathogen sequences also has potential to inform epidemic surveillance and intervention efforts. This is demonstrated clearly in our analysis whereby our results show a much narrower uncertainty and more realistic estimates compared to sampling from the prior. Indeed, the added value of fitting to only 20 local sequences in this analysis demonstrates the utility of phylodynamic modelling for outbreaks as compared to traditional epidemiological modelling fitted only to case data.

It is also worth noting that the analysis described in this report was accomplished in approximately 48 hours and drew on previously developed models and packages for BEAST2 (***Bouckaert et al., 2019; Volz and Siveroni, 2018***). It is therefore feasible for phylodynamic analysis to provide a rapid supplement to epidemiological surveillance, however this requires rapid sequencing and timely sharing of data as well as randomised concentrated sampling of the epidemic within localities such as individual cities.

## Data Availability

Sequences are deposited in GISAID.

https://github.com/emvolz/weifang-sarscov2

https://www.gisaid.org/

## Funding

This work was supported by Centre funding from the UK Medical Research Council (MRC) under a concordat with the UK Department for International Development. NIHR.J-IDEA. Fundingwasalso provided by the MRC Doctoral Training Partnership studentship. This work was also supported by a grant from the Special Project for Prevention and Control of Pneumonia of New Coronavirus Infection in Weifang Science and Technology Development Plan in 2020 (2020YQFK015) to Associate Senior Technologist Qing Nie. Role of the Funders: All funders of the study had no role in study design, data analysis, data interpretation, or writing of the report.

## Acknowledgements

We gratefully acknowledge China National GeneBank at Shenzhen, China for the sequencing strategy and capacity support. We also gratefully acknowledge the laboratories that have contributed publicly available genomes via GISAID: Shanghai Public Health Clinical Center&School of Public Health, Fudan University, Shanghai, China, at the National Institute for Viral Disease Control and Prevention, China CDC, Beijing, China, at the Institute of Pathogen Biology, Chinese Academy of Medical Sciences & Peking Union Medical College, Beijing, China, at the Wuhan Institute of Virology, Chinese Academy of Sciences, Wuhan, China, at the Department of Microbiology, Zhejiang Provincial Center for Disease Control and Prevention, Hangzhou, China, at the Guangdong Provincial Center for Diseases Control and Prevention at the Department of Medical Sciences, at the Shenzhen Key Laboratory of Pathogen and Immunity, Shenzhen, China, at the Hangzhou Center for Disease and Control Microbiology Lab, Zhejiang, China, at the National Institute of Health, Nonthaburi, Thailand, at the National Institute of Infectious Diseases, Tokyo, Japan, at the Korea Centers for Disease Control & Prevention, Cheongju, Korea, at the National Public Health Laboratory, Singapore, at the US Centers for Disease Control and Prevention, Atlanta, USA, at the Institut Pasteur, Paris, France, at the Respiratory Virus Unit, Microbiology Services Colindale, Public Health England, and at the Department of Virology, University of Helsinki and Helsinki University Hospital, Helsinki, Finland, and at the University of Melbourne, Peter Doherty Institute for Infection and Immunity, Melbourne, Australia, at the Victorian Infectious Disease Reference Laboratory, Melbourne, Australia, at the Public Health Virology Laboratory, Brisbane, Australia and at the Institute of Clinical Pathology and Medical Research, University of Sydney, Westmead, Australia.

## Data availability

Genetic sequence data are available from GISAID (gisaid.org). Accession numbers:

EPI_ISL_413691 EPI_ISL_413693 EPI_ISL_413694

EPI_ISL_413695 EPI_ISL_413696 EPI_ISL_413697

EPI_ISL_413711 EPI_ISL_413729 EPI_ISL_413746

EPI_ISL_413747 EPI_ISL_413748 EPI_ISL_413749

EPI_ISL_413750 EPI_ISL_413751 EPI_ISL_413752

EPI_ISL_413753 EPI_ISL_413761 EPI_ISL_413791

EPI_ISL_413809 EPI_ISL_413692 EPI_ISL_404253

EPI_ISL_406717 EPI_ISL_407976 EPI_ISL_412979

EPI_ISL_413854 EPI_ISL_406593 EPI_ISL_408511

EPI_ISL_410301 EPI_ISL_408484 EPI_ISL_406531

EPI_ISL_411066 EPI_ISL_410720 EPI_ISL_411915

EPI_ISL_406862 EPI_ISL_409067 EPI_ISL_413853

EPI_ISL_408666 EPI_ISL_413861 EPI_ISL_414510

EPI_ISL_410536 EPI_ISL_413855 EPI_ISL_413864

EPI_ISL_412965 EPI_ISL_414479 EPI_ISL_414481

EPI_ISL_413996 EPI_ISL_414497 EPI_ISL_413555

EPI_ISL_413586 EPI_ISL_413016 EPI_ISL_414574

EPI_ISL_413024 EPI_ISL_413648 EPI_ISL_414025

EPI_ISL_413566 EPI_ISL_414431 EPI_ISL_413489

EPI_ISL_414449 EPI_ISL_414433 EPI_ISL_414563

EPI_ISL_414466 EPI_ISL_414464 EPI_ISL_414548

EPI_ISL_414467 EPI_ISL_414368 EPI_ISL_414468

EPI_ISL_414455 EPI_ISL_414533 EPI_ISL_414552

EPI_ISL_414590

## Appendix 1

In our analysis, we assumed a prior mean of 500 for initial susceptible, which reflected a reasonable belief of the number of susceptible individuals at the beginning of an outbreak producing 44 confirmed cases. We performed a sensitivity analysis on this parameter, changing it from S=500 to S=9,086,241, the latter reflecting the total population of Weifang. Appendix 1 Figure 1 shows that the median estimated cumulative infections is smaller than the reported number of cases. This is an unrealistic posterior trajectory, highlighting the impossibility of such a high S prior. Further, this adds weight to our conclusion that R(t) fell over time;this would only be possible with a smaller initial susceptible.

**Appendix 1 Figure 1.**
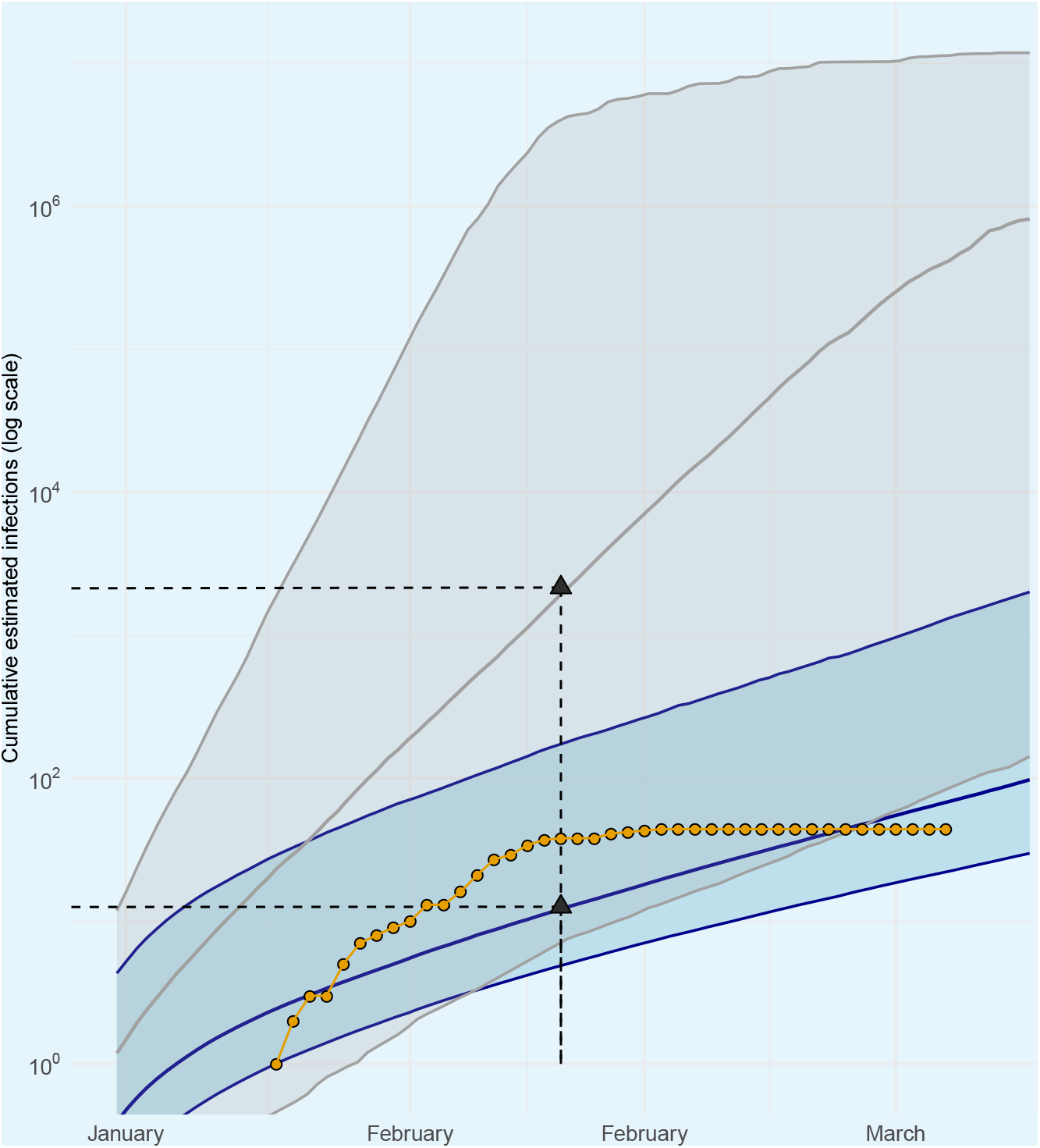
Assuming a mean initial susceptible prior of S = 9,086,241, cumulative estimated infections through time is shown when fitting the SEIR model to genetic data (blue) and sampling only from prior (grey). Solid lines and shaded area reflect posterior median and 95% HPD. The vertical dashed line represents the date of the last sequence sampled in Weifang. Cumulative cases (yellow points) reported by Weifang CDC.

**Appendix 1 Table 1.** Summary of primary epidemiological parameters, including mean estimated posterior and effective sample size due to auto-correlation.

| Statistic | Mean | ESS |
| --- | --- | --- |
| Posterior | -41629.434 | 2998 |
| Likelihood | -41648.33608 | 1057 |
| Prior | 18.90207788 | 1671 |
| Tree likelihood | -41648.33608 | 1057 |
| Tree height | 0.225189455 | 3201 |
| Molecular clock rate | 0.001205673 | 3522 |
| Gamma shape | 0.319969638 | 3174 |
| Transition/transversion | 7.199900029 | 13372 |
| PhydynSEIR | 40.04654004 | 1827 |
| Initial exposed | 4.367728901 | 2403 |
| Initial susceptible | 321.6820578 | 9520 |
| Transmission rate | 21.70707092 | 2757 |
| Initial exogenous | 0.371028398 | 1932 |
| Exogenous growth rate | 19.42898124 | 2156 |
| Migration rate | 1.532603681 | 8091 |

**Figure 1-Figure supplement 1.**
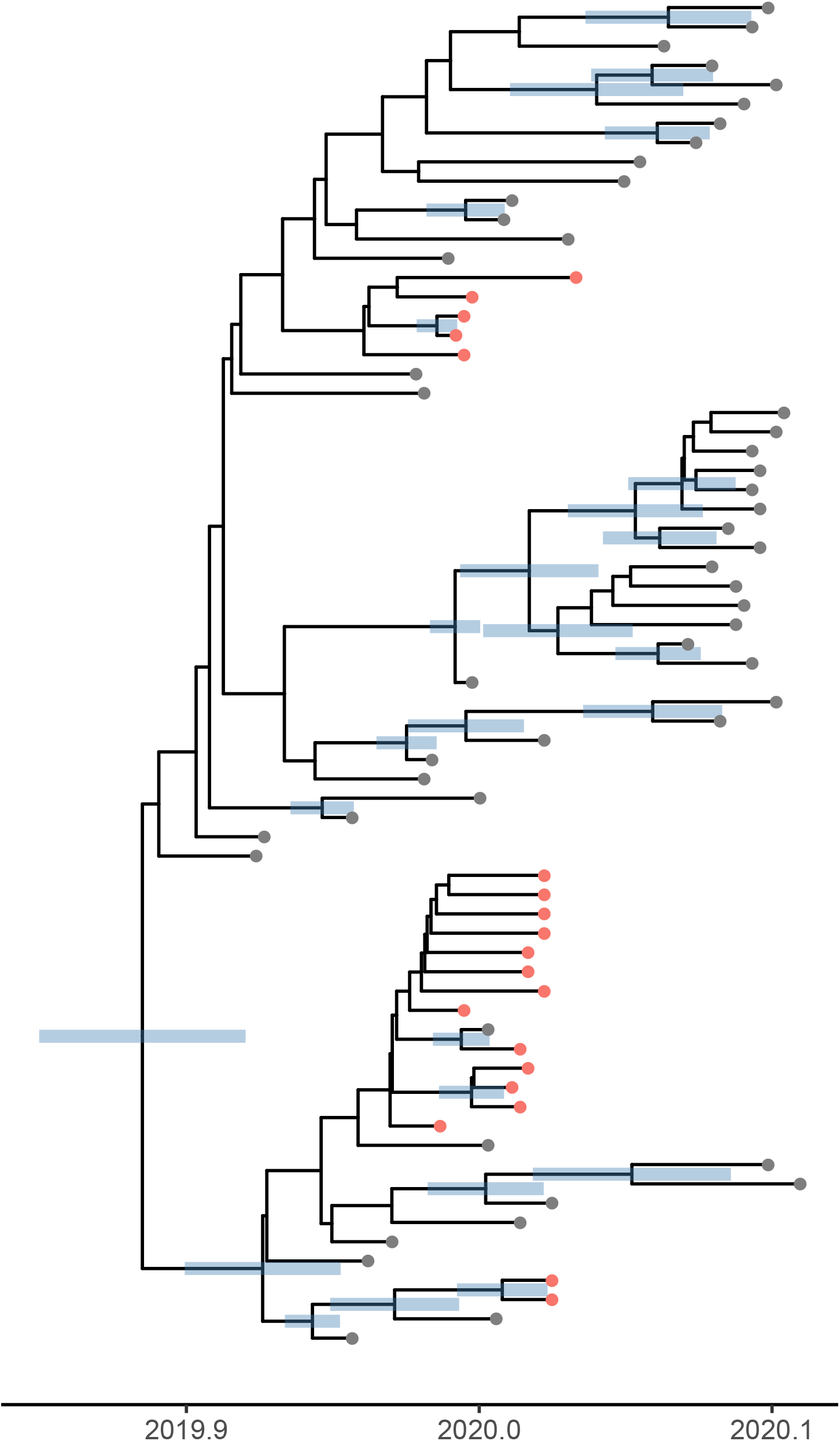
A time scaled phylogeny estimated using IQTree and treedater and using the same data as used for the Bayesian analysis.

**Figure 1-Figure supplement 2.**
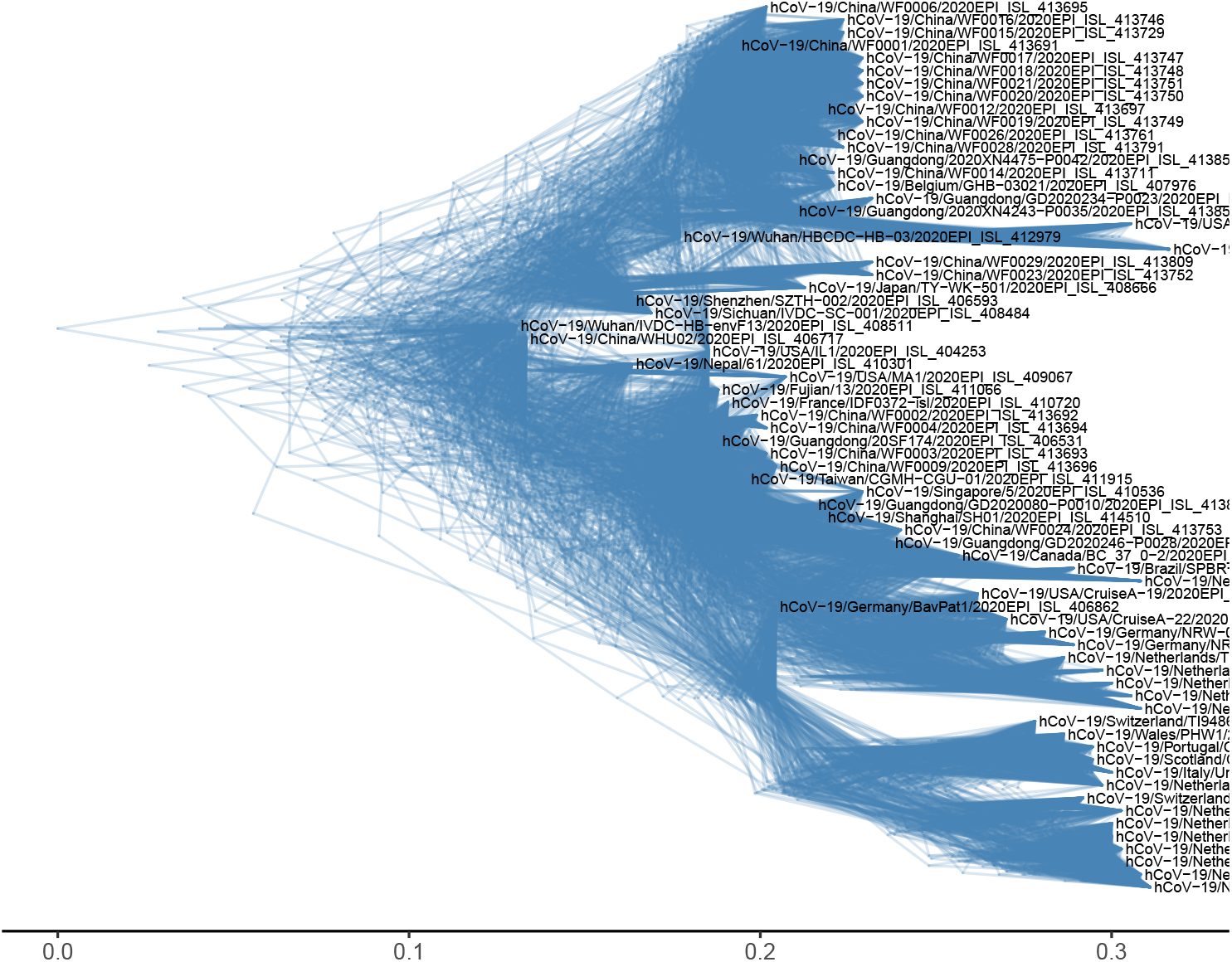
A tree density plot based on the posterior distribution of trees computed in BEAST2.

**Figure 1-Figure supplement 3.**
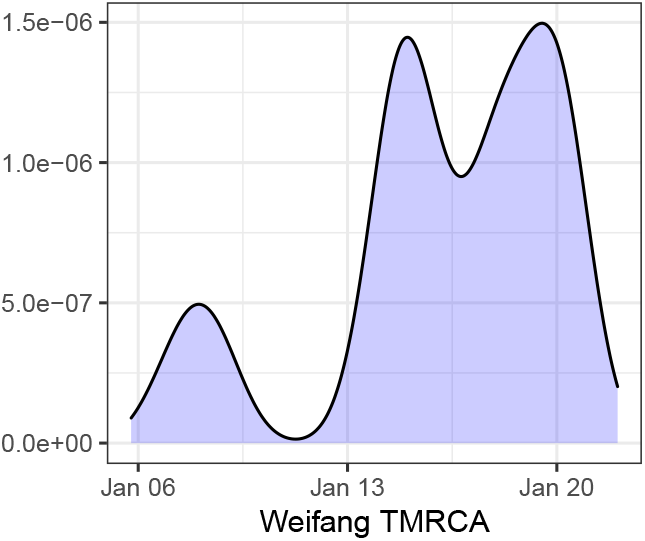
The estimated posterior TMRCA among all Weifang lineages.

**Figure 2-Figure supplement 1.**
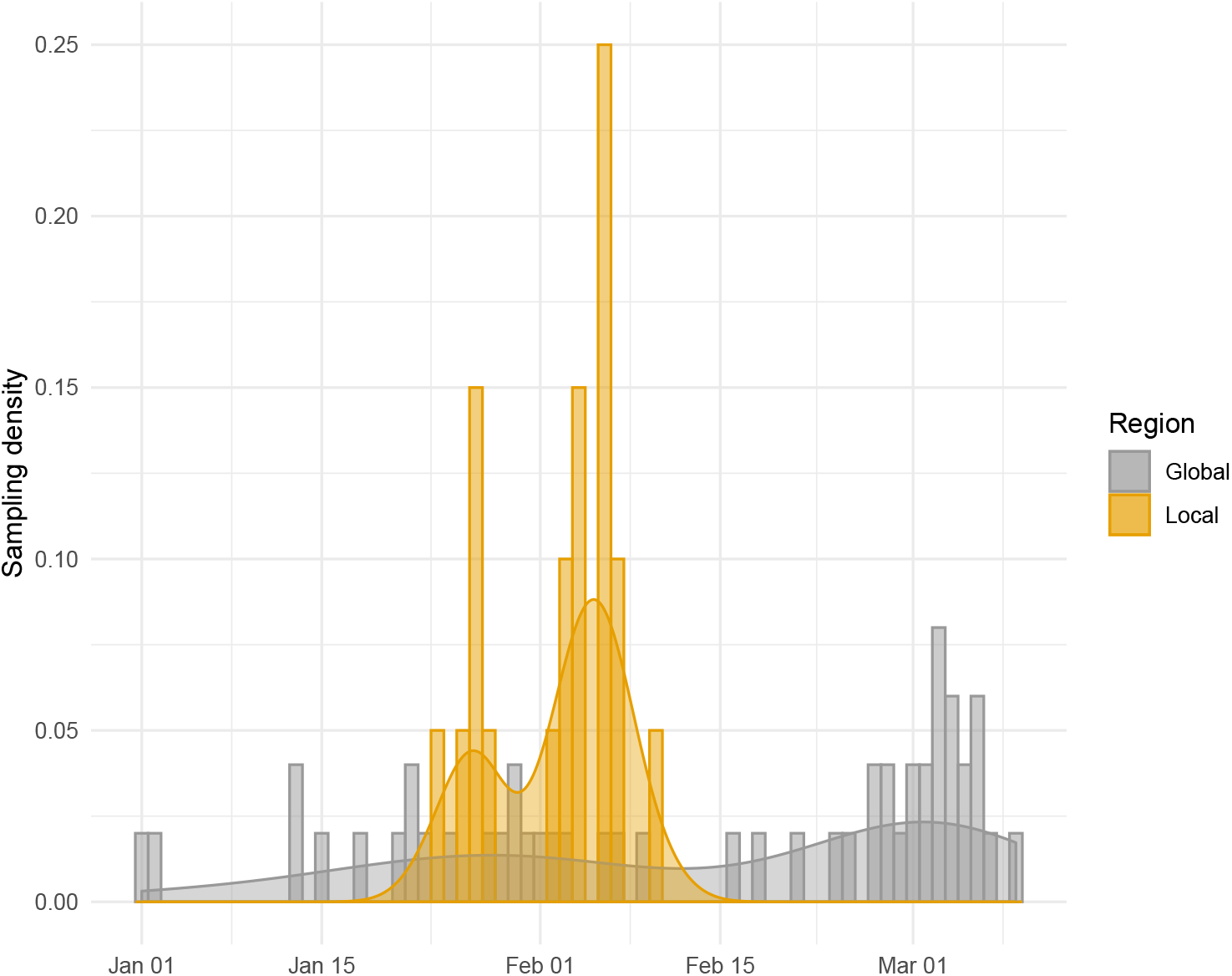
A sample density plot through time of samples inside (yellow) and outside (grey) of Weifang

**Figure 2-Figure supplement 2.**
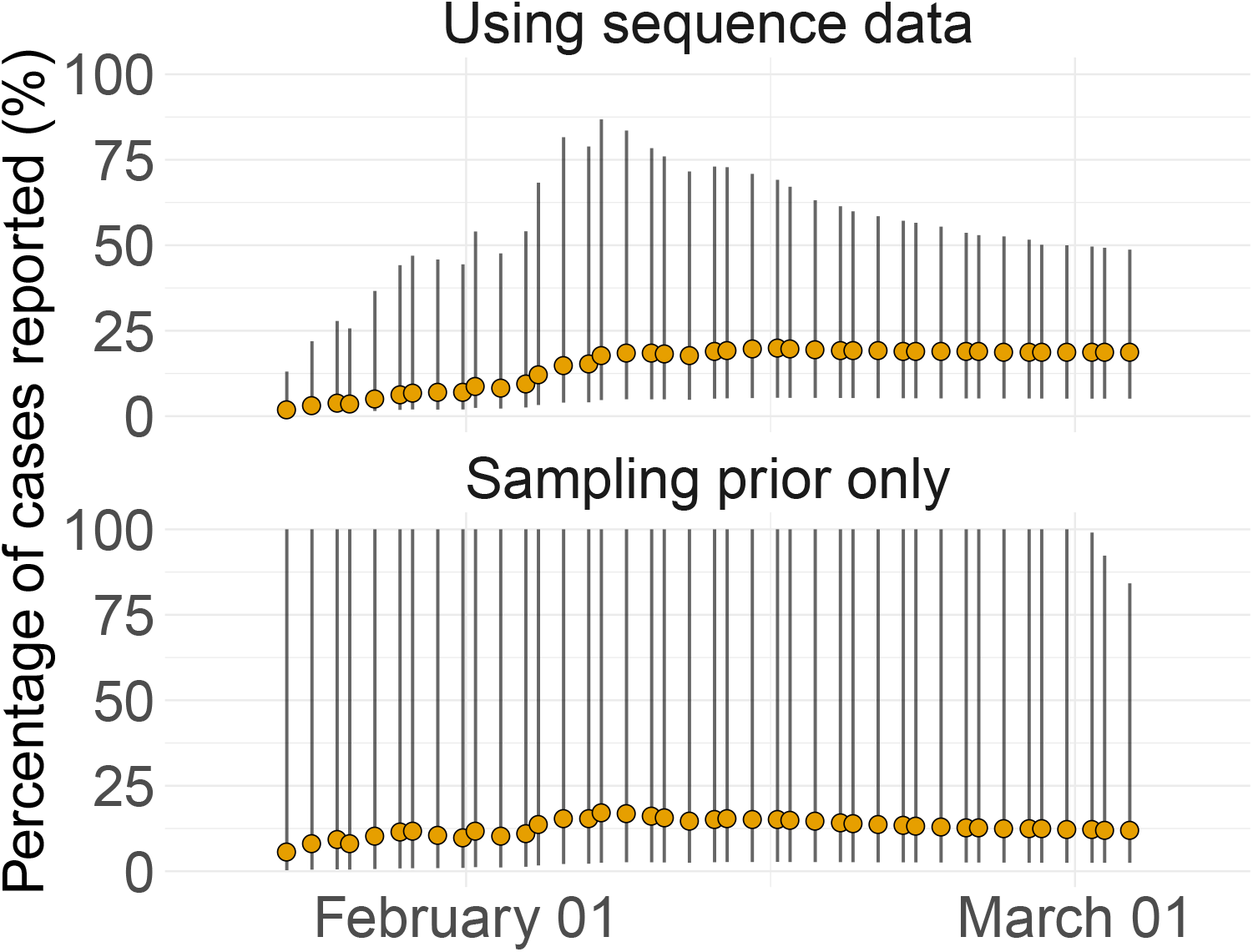
The yellow points and grey bars reflect the mean and 95% HPD cumulative estimated proportion of cases that were identified through time respectively. Results shown separately for analyses conducted with and without the sequence data.

## Notes

### Competing Interest Statement

The authors have declared no competing interest.

### Funding Statement

This work was supported by Centre funding from the UK Medical Research Council under a concordat with the UK Department for International Development. NIHR. J-IDEA. This work was also supported by a grant from the Special Project for Prevention and Control of Pneumonia of New Coronavirus Infection in Weifang Science and Technology Development Plan in 2020 (2020YQFK015) to Associate Senior Technologist Qing Nie. Role of the Funders: All funders of the study had no role in study design, data analysis, data interpretation, or writing of the report.

### Summary of Updates

Re-analysis, sensitivity analysis

